# A Mixed Probiotic/Prebiotic Intervention (MBR-01) for the Management of Diarrhea During Abemaciclib Treatment of Early Breast Cancer: A Single-Center Prospective Case–Control Pilot Study

**DOI:** 10.64898/2026.02.13.26346277

**Authors:** Daniele Generali, Alexandro Membrino, Alessandra Fontana, Federica Gattazzo, Carla Strina, Manuela Milani, Valeria Cervoni, Aldo Caltavituro, Andrea Castagnetti, Sergio Del Bianco, Francesco Schettini

**Affiliations:** Multidisciplinary Unit of Breast Pathology and Translational Research, ASST of Cremona, Cremona, Italy; Department of Medicine, Surgery and Health Sciences, University of Trieste, Cattinara Hospital, 34128 Trieste, Italy; Department for Sustainable Food Process-DiSTAS, Università Cattolica del Sacro Cuore, Piacenza, Italy; Clinical and Translational Oncology, Scuola Superiore Meridionale, Naples, Italy; Translational Genomics and Targeted Therapies in Solid Tumors, August Pi i Sunyer Biomedical Research Institute (IDIBAPS), Barcelona, Spain; Wellmicro srl, via Bruno Rizzi 1, 37012 Bussolengo (VR), Italy; Precision Oncology Department, Sanatrix Clinic, Rome, Italy; Clinic Barcelona Comprehensive Cancer Center, Hospital Clinic Barcelona, Barcelona, Spain

**Keywords:** probiotics, prebiotics, microbiome, abemaciclib, diarrhea, breast cancer

## Abstract

**Background:** Adjuvant abemaciclib+endocrine therapy (ET) improves long-term outcomes in high-risk, hormone receptor–positive (HR+)/HER2-negative early breast cancer (eBC). However, treatment is frequently complicated by diarrhea, affecting adherence and quality of life (QoL). Increasing evidence suggests that abemaciclib-induced gastrointestinal toxicity may involve gut microbiota alterations. We conducted a prospective case-control pilot study evaluating the efficacy of MBR-01, a standardized prebiotic/probiotic formulation, in mitigating abemaciclib-induced diarrhea.

**Methods:** We enrolled 20 patients with high-risk HR+/HER2-negative eBC considered unfit for adjuvant chemotherapy. Patients received abemaciclib+letrozole (control, n=10) or abemaciclib+letrozole+MBR-01 (experimental, n=10). The primary endpoint was the incidence and severity of diarrhea; secondary endpoints included treatment adherence, QoL assessments and exploratory baseline/week-12 microbiota characterization according to treatment arm. Trial registration number: ISRCTN11948182.

**Results:** Diarrhea occurred in all patients. In the control group, diarrhea was predominantly grade 1 (50%) or grade 2 (40%), with one grade 3 event (10%). In the MBR-01 group, diarrhea frequency and severity were reduced by ∼70% at the end of week-12; 80% of patients experienced only grade 1 diarrhea or none by week-12, and no grade ≥3 events. Dose modification was only required in one control. Alpha-diversity and depletion of *F.prausnitzii* were associated with earlier diarrhea onset and longer duration; enrichment in *E.coli* correlated with higher grade events. MBR-01 supplementation seemed to preserve microbial diversity and limited *E.coli* expansion. QoL was significantly improved with MBR-01.

**Conclusion:** MBR-01 may effectively mitigate abemaciclib-induced diarrhea, likely through the achievement of stabilization of gut microbiota composition. Larger prospective studies are warranted to validate these preliminary findings.

**Highlights:**

1. MBR-01, a prebiotic/probiotic, was given to reduce abemaciclib-induced diarrhea.
2. MBR-01 reduced diarrhea by ∼70%, most patients had G0-1, one G ≥3 at week 12.
3. MBR-01 patients keep abemaciclib drug dose; 10% of controls required reduction.
4. MBR-01 halved stool frequency and improved quality of life.
5. MBR-01 preserved gut diversity, maintaining *F. prausnitzii* and limiting *E. coli*.

## Introduction

Hormone receptor–positive (HR+), human epidermal growth factor receptor 2-negative (HER2-) breast cancer represents the most common breast tumor subtype^1^. Adjuvant endocrine therapy (ET) has significantly reduced recurrence risk and mortality in this group. Still, patients with high-risk features continue to experience substantial rates of relapse^2^.

Cyclin-dependent kinase 4 and 6 (CDK4/6) inhibitors have transformed the management of advanced HR+/HER2-breast cancer by prolonging progression-free (PFS) and overall survival (OS) in combination with ET^3,4^. Building on this success, their role in the adjuvant setting has been actively explored. Among available agents, abemaciclib is unique for its continuous dosing schedule and broader kinase inhibition profile^5^. The pivotal monarchE trial demonstrated that adding abemaciclib to ET significantly improved invasive disease-free survival (iDFS) and OS in patients with high-risk early breast cancer^6,7^. On the basis of these results, abemaciclib has become the first CDK4/6 inhibitor (CDK4/6i) approved for adjuvant use. However, the clinical benefits of abemaciclib are tempered by its distinct toxicity profile. Diarrhea occurs in up to 80% of treated patients, with approximately 10–20% experiencing grade ≥3 events^8,9^. While usually manageable with antidiarrheal therapy and dose interruptions or reductions, gastrointestinal toxicity remains the leading cause of treatment modification and can significantly impair adherence and quality of life (QoL). Maintaining dose intensity is critical, given post hoc analyses of monarchE suggesting a correlation between treatment adherence and clinical benefit^10^. The mechanisms underlying abemaciclib-induced diarrhea are not fully understood, but off-target activity on cyclin-dependent kinase 9 (CDK9), glycogen synthase kinase-3β (GSK3β), and calcium/calmodulin-dependent protein kinase II (CAMKII) have been hypothesized to disrupt the balance between epithelial proliferation and differentiation, compromising barrier function and promoting diarrhea^11–13^.

In parallel, research has increasingly highlighted the role of the gut microbiota in modulating treatment efficacy and toxicity across different solid tumors. In fact, gut microbiota is central to immune regulation, drug metabolism, and epithelial integrity^14^, and dysbiosis, characterized by loss of microbial diversity and depletion of protective taxa, has been linked to systemic inflammation, carcinogenesis, and altered responses to systemic therapy^15,16^, including CDK4/6i+ET^17^.

Emerging evidence suggests that abemaciclib interact with the gut microbiota, differently from other CDK4/6i. Indeed, a recent prospective study showed that abemaciclib treatment, but not ribociclib, was associated with reduced alpha-diversity and distinct taxonomic shifts, which may contribute to diarrhea onset and severity^18^.

Probiotics, prebiotics, and postbiotics have shown potential to enhance barrier function, reduce mucosal inflammation, and stabilize microbial ecosystems ^19^. De Sanctis et al. showed promising results in controlling abemaciclib-induced diarrhea with a postbiotic microbiota stabilizer^20^, supporting the concept that microbiota-targeted interventions may help mitigating this relevant side effect. Building on these findings, we designed a prospective pilot study to evaluate whether MBR-01, a standardized probiotic + prebiotic formulation, could reduce diarrhea in patients receiving adjuvant abemaciclib + ET for high-risk early breast cancer and possible correlations between baseline microbiota composition and diarrhea occurrence. The protocol included a butyrate-producing probiotic, the *Clostridium Butyrricum*, which may enhance gut mucosal health, water and electrolyte absorption, and intestinal barrier integrity^21,22^, together with a second probiotic formulation providing *Enterococcus faecium L3*, whose bacteriocins inhibit enteric pathogens and strengthen epithelial barrier function^23,24^, and *Bifidobacterium animalis subsp. lactis BB-1*2, which reinforces mucosal integrity and supports microbiota recovery after dysbiosis^25^. These probiotics were complemented by a prebiotic fibre mixture that supports beneficial gut microbes and helps maintain intestinal microbiota equilibrium^26^.

This study aims to provide early clinical evidence supporting microbiota-targeted supportive care strategies in oncology and to inform future randomized trials.

## Materials and Methods

### Study design and ethical considerations

This prospective, single-center, case–control pilot study was conducted at the Cremona Hospital (Italy), in accordance with the principles of the Declaration of Helsinki and Good Clinical Practice guidelines. The study protocol was reviewed and approved by the institutional ethics committee (approval number: 34326 - 18/10/2019-EC/ATS Val Padana). All patients provided written informed consent prior to enrollment. The trial was registered in the World Health Organization-recognized registry ISRCTN with study registration number ISRCTN11948182^27^.

### Patient selection

Eligible patients were women aged ≥18 years with histologically confirmed HR+/HER2– early breast cancer at high-risk of recurrence. High-risk was defined as ≥4 positive axillary lymph nodes (N2), or 1–3 positive nodes (N1) with additional features such as tumor size ≥5 cm (T3), histologic grade 3 (G3), or Ki-67 ≥20%. Patients were deemed unsuitable for adjuvant chemotherapy due to comorbidities frailty, or patient preference. Additional inclusion criteria were: Eastern Cooperative Oncology Group (ECOG) performance status 0–2, adequate hematologic and organ function, and ability to provide fecal samples. Exclusion criteria included: prior exposure to CDK4/6i, chronic diarrheal disorders, inflammatory bowel disease, intestinal resection affecting absorption, systemic antibiotic, proton pump inhibitors or probiotic use within 4 weeks prior to enrollment, immunosuppressive treatment, or concurrent participation in another interventional study.

### Treatment groups and interventions

Patients who refused chemotherapy due to the presence of relevant comorbidities such as reduced left ventricular ejection fraction (LVEF) or overall frailty and personal circumstances, including preference to avoid alopecia and the need to maintain work or caregiving responsibilities were enrolled into the study. Patients were assigned to the control or intervention group based on their willingness to receive prophylactic MBR-01, a standardized probiotic and prebiotic protocol designed by the Multidisciplinary Oncology Unit at the Cremona Hospital. Patients in the experimental arm received the MBR-01. The control group received abemaciclib 150 mg twice daily + letrozole 2.5 mg daily. The experimental group received the same regimen together with MBR-01. Treatment was administered for 12 consecutive weeks. No prophylactic anti-diarrheal medication was permitted, although loperamide could be used at the discretion of the treating physician if clinically required or indicated.

### Study endpoints

The primary endpoint was the incidence and severity of abemaciclib-induced diarrhea, graded according to the National Cancer Institute Common Terminology Criteria for Adverse Events (NCI-CTCAE) version 5.0 at week 12. Secondary endpoints included: the need for dose reductions, treatment interruptions or discontinuations due to gastrointestinal toxicity, and patient-reported stool frequency and consistency recorded through daily electronic diaries. Health-related QoL (HRQoL) was also a secondary endpoint and was assessed using the European Organisation for Research and Treatment of Cancer Quality of Life Questionnaire (EORTC QLQ)-C30 and QLQ-BR23, focusing on predefined functional domains from both instruments, namely QLQ-C30’s emotional, role, and physical functioning and QLQ-BR23’s body image and sexual functioning. An exploratory QoL interference score (custom composite) and diary-based stool frequency were analyzed as additional patient reported outcomes (PROMs). An exploratory endpoint assessed changes in gut microbiota composition and diversity from baseline to week 12, together with their potential association with diarrhea occurrence and severity.

### Clinical assessments

Clinical evaluations were performed at baseline and every 28 days. At each visit, the incidence, grade, onset and duration of diarrhea were recorded and cross-checked with daily electronic diaries (care@you®), which also captured patient-reported stool frequency and consistency. Treatment adherence was assessed using pill counts and patient self-report. Laboratory monitoring, including hematology and biochemistry, was performed at each visit according to standard practice.

### Stool sample collection and processing, DNA extraction and purification

Fecal samples were collected at baseline and at day 84 (week 12). Patients were instructed to collect mid-stool aliquots using the standardized collection kits SMART-eNAT^®^ (Copan Italia S.p.a., Brescia, Italy). Samples were immediately stabilized in collection medium, stored at ambient temperature, and transported within 24 hours to the study site, where they were frozen at –80 °C until analysis. Negative controls (blank collection kits) and positive controls (mock microbial community standards) were processed in parallel to monitor potential contamination. Fecal samples were mechanically lysed using inert beads and the FastPrep^®^ homogenizer (MP Biomedicals, Irvine, CA, USA). DNA extraction and purification was performed using the Mag-Bind^®^ Universal Pathogen Kit (Omega Bio-Tek, Norcross, GA, USA) on the automated NIMBUS Presto platform (Hamilton Company, Reno, NV, USA). Microbial DNA was quantified using the Qubit™ 1X dsDNA Broad Range Assay Kit (Thermo Fisher Scientific, Waltham, MA, USA) on the Synergy Instrument (Agilent Technologies, Santa Clara, CA, USA) and normalized to a concentration of 13.3 ng/µl.

### Bioinformatic analysis

Metagenomic libraries were generated with the Illumina^®^ DNA Prep kit (Illumina, San Diego, CA, USA) following the manufacturer’s protocol adapted for the VANTAGE automated platform (Hamilton Company). Samples were subjected to capillary electrophoresis using the Qiaxcel™ instrument (Qiagen, Hilden, Germany). The libraries were quantified with the Qubit™ 1X dsDNA High Sensitivity Assay Kit (Thermo Fisher Scientific) and normalized to a concentration of 4 nM and sequenced on the NovaSeq6000Dx system (Illumina). Quality control of raw reads was performed using FastQC. Taxonomic profiling was carried out using MetaPhlAn4, enabling species-level resolution.

### Statistical analyses

Descriptive statistics were used to summarize baseline characteristics and clinical variables. Proportions between treatment groups were compared using Fisher’s exact test. Continuous variables, including patient-reported outcomes (PROs) and HRQoL scores, were analyzed using the Mann–Whitney U test for between-group comparisons and the Wilcoxon signed-rank test for within-group changes from baseline to week 12. For microbiome analysis, alpha diversity (Shannon, Simpson and Chao1 indices) was compared within and between groups using non-parametric tests as appropriate. Beta diversity was assessed using Bray–Curtis and weighted UniFrac distances and compared by PERMANOVA. Differential taxonomic analyses examined changes between baseline and week 12, and significance was corrected for multiple testing using the false discovery rate (FDR). Correlations between microbial features and clinical variables, including diarrhea onset, duration and grade, were assessed using Spearman’s coefficient and considering only the taxa showing the most pronounced and statistically significant shifts in the overall comparative analysis. A multivariable logistic regression model was used to evaluate whether relevant clinical and microbial baseline features independently predicted the occurrence of grade ≥2 diarrhea. The study was exploratory, with no formal sample size calculation. All statistical tests were two-sided, and p-values <0.05 were reported descriptively to identify preliminary signals of activity of the MBR-01 combination. As the study was purely exploratory, numerical trends were prioritized over formal statistical significance, and no correction for multiplicity was applied. All analyses were performed in R version 4.3.2 for MacOSX.

## Results

### Patient characteristics

Between January 2023 and April 2025, 20 patients with high-risk HR+/HER2– early breast cancer were enrolled: 10 in the control group and 10 in the intervention group (**Table 1**). Baseline demographic and clinical characteristics were similar between the two groups. No significant differences were observed in comorbidities. The overall median age was 72 years (range, 65-79 years), with 100% of patients being postmenopausal, 70% with ECOG performance status 0 and 30% with ECOG performance status 1. As per inclusion criteria, patients had not been exposed to systemic antibiotics, probiotics and proton pump inhibitors before study treatment start.

**Table 1.** Baseline Characteristics of the Study Population.

| VARIABLES | Control | Experimental |
| --- | --- | --- |
|  | N (%) | N (%) |
|  | 10 (100%) | N (100%) |
| Median age, years (min-max) | 73 (68-79) | 68 (65-72) |
| Menopausal status |  |  |
| *Postmenopausal* | 10 (100) | 10 (100) |
| *Pre/perimenopausal* | 0 (0) | 0 (0) |
| ECOG* |  |  |
| 0 | 4 (40) | 8 (80) |
| 1 | 6 (60) | 2 (20) |
| 2 | 0 (0) | 0 (0) |
| T<sup>#</sup> |  |  |
| <5cm | 6 (60) | 7 (70) |
| ≥5 cm | 4 (40) | 3 (30) |
| N<sup>§</sup> |  |  |
| 1-3 positive lymphnodes | 8 (80) | 5 (50) |
| ≥4 positive lymphnodes | 2 (20) | 5 (50) |
| Endocrine therapy |  |  |
| *Aromatase inhibitor* | 10 (100) | 10 (100) |
| *Tamoxifen* | 0 (0) | 0 (0) |

**Footnotes** \*: Fisher's exact test p=0.170; #: Fisher's exact test p= 1.00; §: Fisher's exact test p=0.350.

### Incidence and severity of diarrhea

Diarrhea was reported in all patients receiving abemaciclib in the control group, with five patients (50%) experiencing grade 1, four patients (40%) grade 2 and one patient (10%) grade 3 diarrhea after the first cycle. In the MBR-01 group, 80% patients reported diarrhea after the first cycle, 40% of grade 2. By week 12, 90% of patients in the MBR-01 group reported either grade 1 diarrhea or no diarrhea, compared with 50% in the control group. No patient in the MBR-01 group experienced grade ≥3 diarrhea. Dose reductions due to gastrointestinal toxicity were required in one patient (10%) in the control group, whereas none were needed in the MBR-01 group. All patients in both arms completed the planned 12 weeks of therapy. Although the difference between groups at weeks 12 did not reach statistical significance (p=0.141), the observed trend favored the MBR-01 regimen (**Figure 1**). Median time to diarrhea onset was 8 days (range, 4-14 days), and the median duration of episodes was 9-10 days (interquartile range [IQR], 7 - 12 days) for grade 2 and 6-7 days (IQR 4 - 9 days) for grade 1.

**Figure 1.**
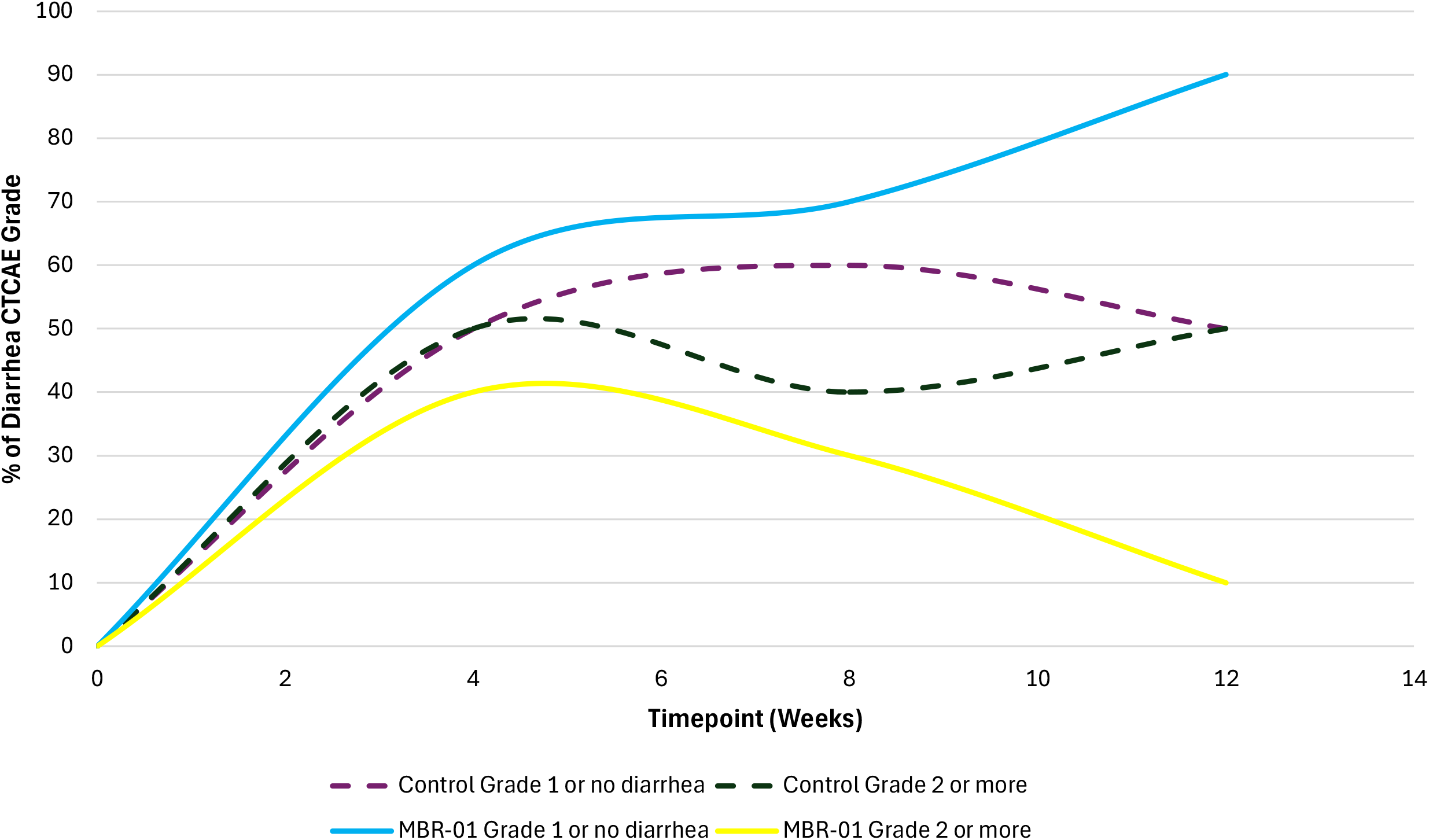
Trend of incidence and severity of diarrhea graded according to CTCAE v5.0 criteria. CTCAE: Common Terminology Criteria for Adverse Events.

### Patient-reported outcomes, QoL and treatment adherence

Daily electronic diaries (care@you^®^) confirmed investigator-reported diarrhea events and provided additional granularity on stool frequency and consistency. Control group patients reported a median of 4.0 (IQR 3.3 – 4.8) stools/day during diarrheal episodes, representing a significant increase (p=0.013) from the pre-treatment median of 1.0 (IQR 1.0 - 1.8) stool/day. Conversely, the experimental group reported a significantly lower median of 2 (IQR 1 - 2.8) stools/day as compared to controls (p=0.022), without any significant increase in frequency (p=0.484) during the 12 weeks of abemaciclib supported by MBR-01 treatment.

By the end of the 12-week treatment, the QoL interference scores were significantly lower in the experimental group as compared to control, with a median score of 2.5 (IQR, 2.0 – 3.0) vs. 4.5 (IQR, 3.0 – 5.8) (p=0.002), with a slight reduction from baseline in the former group (p=0.037) and significant increase in the latter (p=0.014). Sexual functioning (p=0.673) and emotional functioning (p=0.270) scores were not different between groups, as well as the body image score (p=0.545), whilst role functioning score (median 73.5, [IQR 72.0 - 80.0] vs. 61.0, [IQR 59.0 - 63.5]) and physical functioning score (median 80.00, [IQR 71.8 - 84.25] vs. 65.5, [IQR 62.5 - 72.8]) were significantly higher in the experimental arm after 12 weeks vs. controls (p<0.001 and p=0.021), without experiencing a significant modification from baseline (p=0.286 and p=1.00). In the control group, emotional, role and physical functioning scores all decreased significantly after 12 weeks (**Figure 2**). The treatment adherence was numerically higher in the experimental group compared with controls. All patients in the interventional group maintained full abemaciclib dosing throughout the 12-week period, whereas one patient (10%) in the control arm required a dose reduction due to persistent grade 3 diarrhea (p=0.305). No permanent discontinuations occurred in either group.

**Figure 2.**
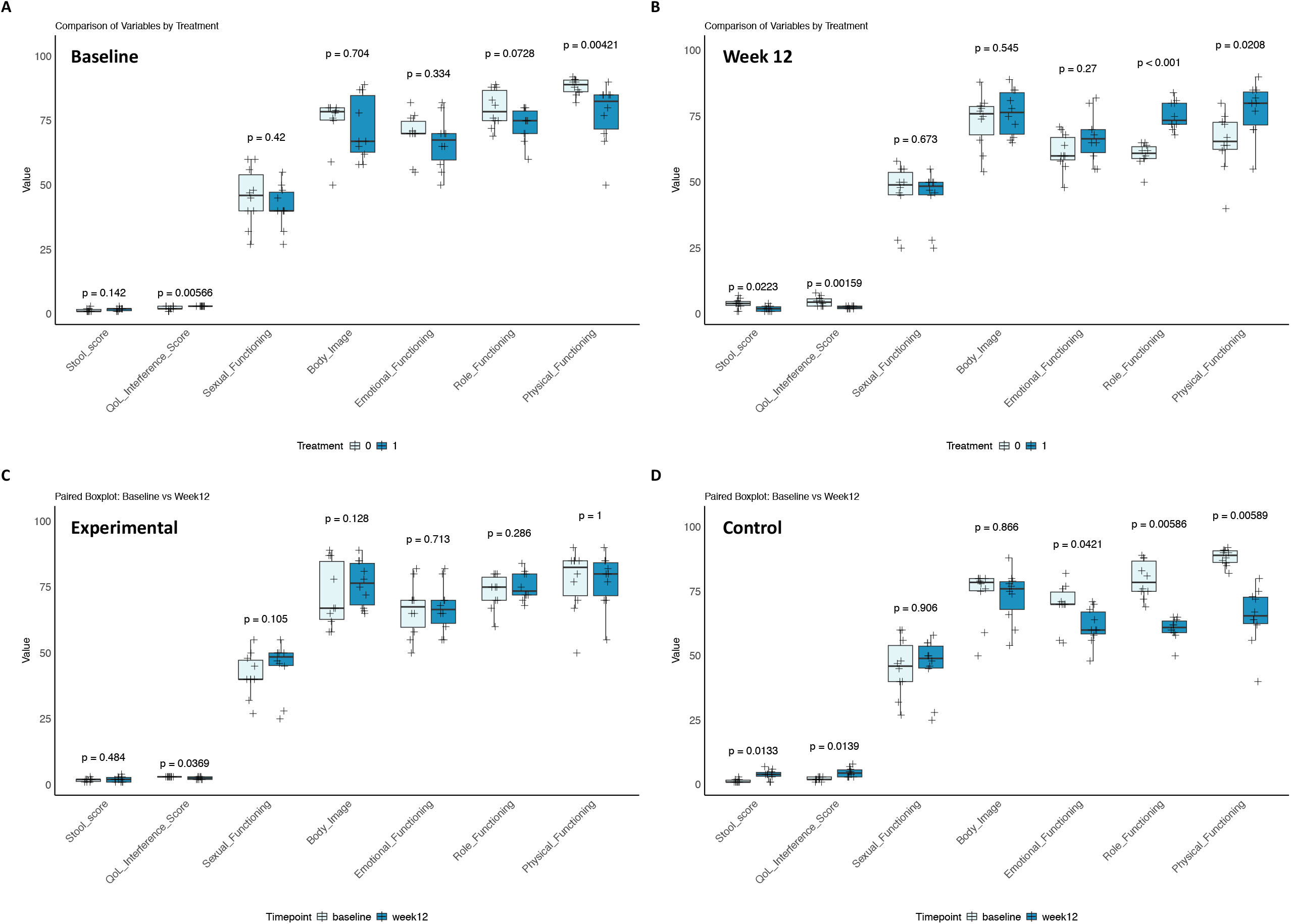
Paired and unpaired comparisons among QoL scores of interest. Scores’ distribution at baseline according to treatment group. **B:** Scores’ distribution at week 12 according to treatment group. **C:** Scores’ distribution at baseline and week 12 in the experimental group. **D:** Scores’ distribution at baseline and week 12 in the control group. P values in A and B are referred to Mann Whitney U tests for unpaired samples, while in C and D are referred to Wilcoxon signed rank tests for paired samples; 0=control arm; 1=experimental arm; QoL=quality of life.

### Exploratory microbiome analysis

At baseline, alpha-diversity indices (Shannon, Simpson, Chao1) were comparable between groups (all p>0.05). Longitudinal analyses focused on the Shannon index, given its robustness and widespread use for capturing within-sample diversity changes. By day 84, patients in the control group exhibited a significant reduction in microbial alpha-diversity (median Shannon index 3.7 at baseline vs. 2.7 at day 84, p=0.010). In contrast, MBR-01 supplementation preserved microbial diversity, with only a modest, non-significant decline observed (median Shannon index 3.8 vs. 3.5, p=0.210) (**Figure 3A**). Beta-diversity analysis using Bray-Curtis dissimilarity and weighted UniFrac showed no significant differences between groups at baseline (PERMANOVA p>0.05), confirming comparable starting microbial compositions. By week 12, distinct clustering of microbial communities was observed between control and MBR-01 groups (PERMANOVA p=0.030), suggesting that MBR-01 modulated the trajectory of abemaciclib-associated microbial shifts.

**Figure 3.**
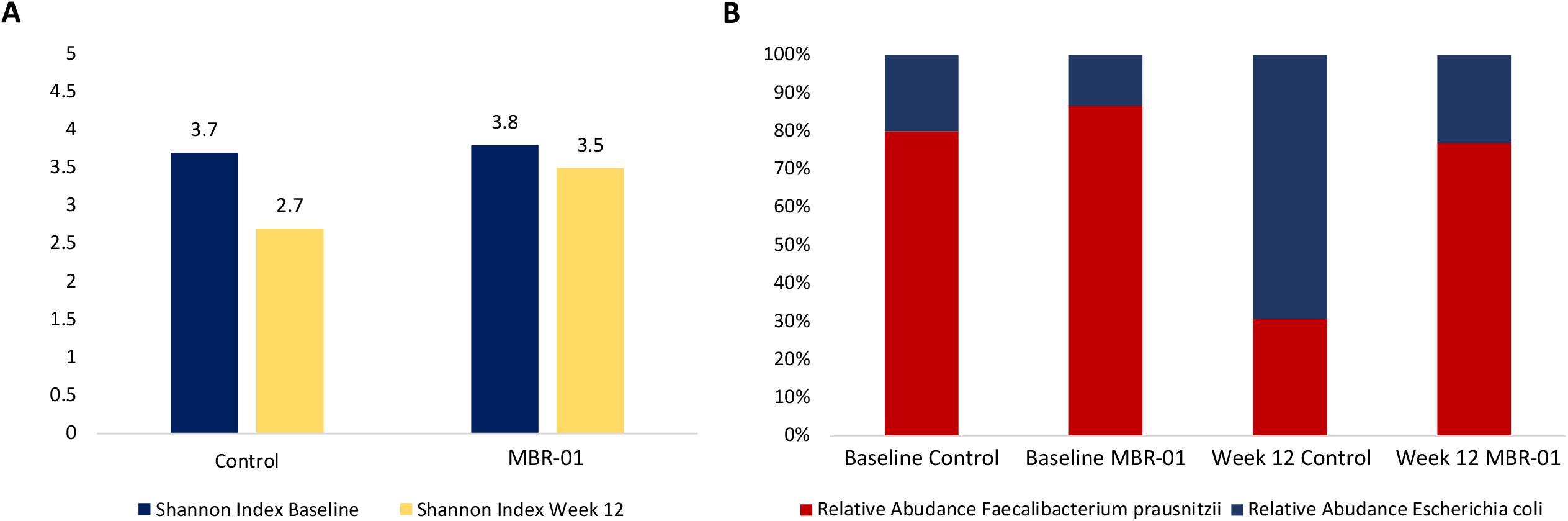
Alpha diversity and taxonomic shifts in selected fecal microbiota key taxa. Shannon Index median values (alpha diversity) at baseline and week 12 for control and MBR-01 groups. Higher values indicate greater microbial diversity. **B**: relative abundance of *Faecalibacterium prausnitzii* (red) and *Escherichia coli* (blue), expressed as normalized proportions of total reads. Data illustrate a reduction in diversity and beneficial taxa in the Control group over time, while MBR-01 maintains a more stable microbial profile.

### Taxonomic shifts associated with diarrhea

Taxonomic profiling revealed significant shifts in gut microbiota composition after 12 weeks (FDR<0.05). The taxa with the most pronounced changes between baseline and week 12 were *Faecalibacterium prausnitzii* and *Escherichia coli*, providing the rationale for subsequent focus. In the control group, *Faecalibacterium prausnitzii* declined markedly from a mean of 12% at baseline to 4% at week 12 (p<0.010). Concurrently, *Escherichia coli* expanded from 3% to 9% (p=0.040), correlating with diarrhea onset and severity (**Figure 3B**). Correlation analyses showed that lower baseline *Faecalibacterium prausnitzii* abundance correlated with earlier diarrhea onset (Spearman r =-0.62, p=0.020) and longer episode duration, while *Escherichia coli* enrichment correlated with higher maximum diarrhea grade (r=0.55, p=0.040). In the MBR-01 group, *Faecalibacterium prausnitzii* levels were preserved (13% at baseline vs. 10% at week 12, p=0.210), and no significant *Escherichia coli* expansion was observed (2% to 3%, p=0.180) (**Figure 3B**).

### Clinical–microbiota correlations

Multivariate logistic regression incorporating baseline microbial diversity, age, body mass index (BMI), ECOG performance status, and treatment group identified two independent predictors of grade ≥2 diarrhea: the reduced microbial alpha-diversity (OR 3.8, 95% CI 1.2–11.5, p=0.02) and low baseline abundance of *Faecalibacterium* (OR 4.1, 95% CI 1.3–12.8, p=0.01), while the MBR-01 intervention was independently associated with a reduced risk of grade ≥2 diarrhea (OR 0.25, 95% CI 0.07–0.88, p=0.03). No significant associations were observed for age, BMI, or ECOG status (**Figure 4**).

**Figure 4.**
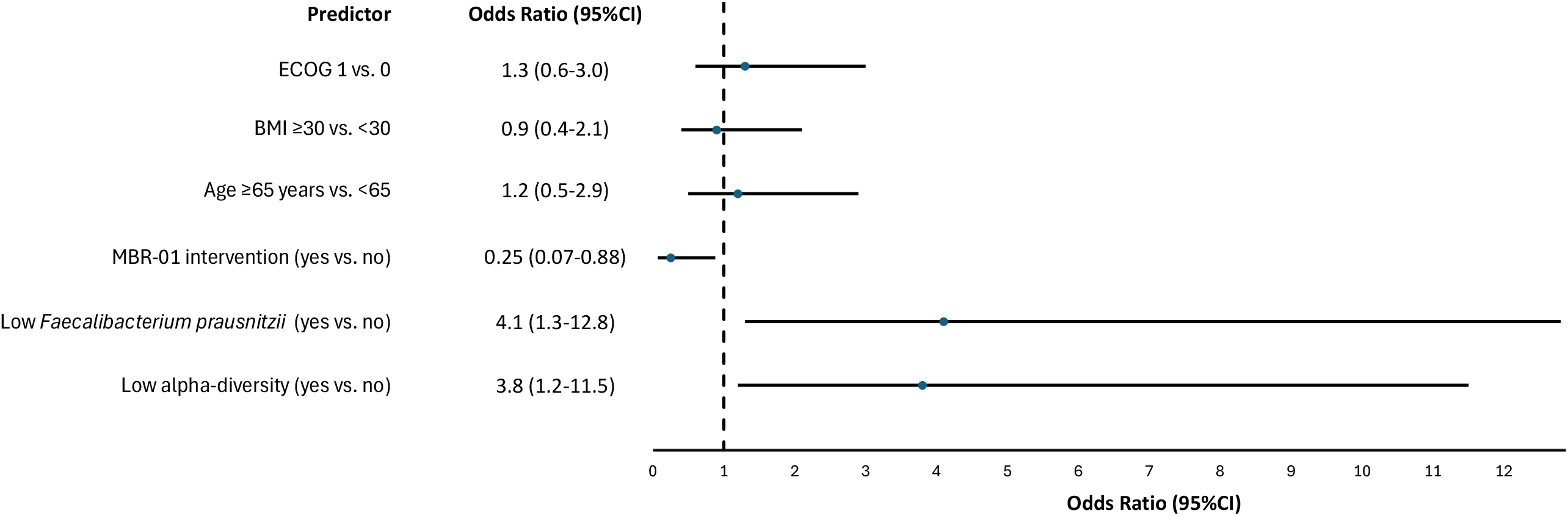
Multivariable analysis of clinical–microbiota associations with reduced risk of grade ≥2 diarrhea. Forest plot of multivariable logistic regression including microbiota alfa-diversity at baseline and clinical patients characteristics. Cut-off for low alpha-diversity was Shannon index<3.5. Cut-off for low relative abundance of *Faecalibacterium* <5%. BMI: body mass index; CI: confidence intervals.

## Discussion

This prospective, single-center pilot study provides preliminary evidence that MBR-01, a standardized probiotic/prebiotic formulation, may mitigate abemaciclib-induced diarrhea, likely preserving gut microbiota diversity in patients with high-risk HR+/HER2-early breast cancer. Adjuvant abemaciclib in combination with ET represents a significant advance for patients with high-risk HR+/HER2– early breast cancer^6,7^. However, its therapeutic benefit is counterbalanced by a high incidence of diarrhea, including grade 3 or higher in 1/5 of the cases^8,9^. Our study confirmed that diarrhea was nearly universal among abemaciclib-treated patients. Although generally manageable with antidiarrheal medications and dose modifications, diarrhea is the most common reason for treatment interruptions and can negatively impact adherence, QoL, and potentially, treatment efficacy^10^. In fact, in our study, the control group experienced a significant decline in emotional, role and physical functioning, alongside an increased stool deposition frequency and a clear interference with their QoL, with 10% patients requiring abemaciclib dose reduction. None of this happened in the experimental group where, in turn, a slight reduction of the QoL interference score was observed alongside reduction in diarrhea CTCAE grade and preservation of abemaciclib full dose, with MBR-01 supplementation. Notably, no grade ≥3 diarrhea occurred in the MBR-01 group. These findings suggest that microbiota-targeted strategies may help overcome one of the most clinically relevant barriers to sustained abemaciclib exposure.

The pathogenesis of abemaciclib-induced diarrhea is multifactorial. Preclinical studies have shown abemaciclib uniquely causes crypt hyperplasia, villous blunting, and mucosal inflammation in rodent models^13^. These types of changes have not been observed with palbociclib or ribociclib. In addition to CDK4/6 inhibition, abemaciclib targets CDK9, GSK3β, and CAMKII, integral kinases of the intestinal epithelial homeostasis^12^. Disruption of these pathways likely contributes to epithelial barrier dysfunction and increased susceptibility to diarrhea. Our microbiota analysis further supports the hypothesis that gut dysbiosis amplifies gastrointestinal toxicity. Control patients demonstrated a significant decline in alpha-diversity during abemaciclib therapy, alongside depletion of *Faecalibacterium prausnitzii*, a key butyrate producer essential for barrier integrity^28,29^, and expansion of *Escherichia coli*, a pathobiont often associated with inflammation^30^. Both features correlated with earlier onset, longer duration, and higher severity of diarrhea. Conversely, MBR-01 supplementation preserved microbial diversity, seemed to attenuate *Faecalibacterium* loss and limit *Escherichia coli* overgrowth, suggesting that stabilization of microbial ecosystems may be protective against toxicity. These findings are consistent with prior reports linking microbiota diversity to resilience against treatment-related toxicity^16^. Together, these data reinforce the notion that drug–microbiota interactions are bidirectional, i.e. abemaciclib perturbs the microbiota, which in turn may exacerbate gastrointestinal toxicity.

Conventional strategies for managing abemaciclib-induced diarrhea, mainly loperamide and dose modifications, are often reactive and do not address underlying biological mechanisms. Our results provide preliminary evidence that a mixed probiotic/prebiotic intervention can reduce gastrointestinal toxicity while favorably modulating microbiota composition. Mechanistically, prebiotics may act through multiple, complementary pathways: restoration of epithelial barrier integrity, stimulation of short-chain fatty acid (SCFA) production (notably butyrate), suppression of pro-inflammatory signalling, and ecological stabilization of microbial communities^31,32^. Clinical data support a heterogeneous picture in practice. A recent pragmatic 2-group study using a commercially-available postbiotic reported fewer severe diarrhea events and fewer dose reductions during abemaciclib therapy compared with historical controls, suggesting benefit from microbiota stabilizers in this setting^20^. By contrast, the randomized MERMAID trial testing a probiotic (*Bifidobacterium*) ± trimebutine maleate did not reduce the incidence of grade ≥2 diarrhea compared with historical control, although grade 3 events were infrequent and some reduction in highest-grade events was observed, underscoring that not all microbiota-modulating strategies produce the same clinical effect^33^. Taken together, these data suggest two practical implications: first, supplementation with a defined combination of probiotics (e.g., *Clostridium butyricum, Enterococcus faecium L3* and *Bifidobacterium animalis subsp. lactis BB-12*) together with complementary prebiotic fibers, as done in our study, may provide broader translational benefits than single-strain probiotics, by promoting gut microbial community balance, enhancing SCFA production, and supporting mucosal barrier function; second, a precision approach in selecting live-microbe products, tailoring probiotic or symbiotic choices to an individual’s baseline microbiota, is biologically plausible and may increase the likelihood of clinical benefit.

Our study has several limitations: 1) it is a pilot study designed to detect preliminary signals of efficacy, without power to draw any definitive conclusion; 2) the sample size was small and the study was conducted at a single center, limiting generalizability; 3) the intervention was not randomized, introducing potential selection bias; 4) follow-up was limited to 12 weeks, and longitudinal changes in microbiota beyond this timeframe remain unknown. Finally, while correlations between microbial features and diarrhea were observed, causality cannot be definitively established. At the same time, our findings have several clinical implications worthy of further exploration. Firstly, microbiota-targeted prophylaxis with MBR-01 reduced diarrhea severity and frequency, potentially improving adherence to abemaciclib, a factor directly associated with therapeutic benefit in the adjuvant setting^10^. Secondly, baseline microbial features such as low alpha-diversity and reduced *Faecalibacterium* abundance in patients experiencing more diarrhea onset and higher grade, suggest that microbiota profiling could help identify patients most likely to benefit from prophylactic interventions. Thirdly, the increase in *Escherichia coli* in the group experiencing more diarrhea suggests the potential to dynamically monitoring fecal microbiota composition to proactively detect patients without baseline dysbiosis who could develop it during treatment and may potentially benefit from the introduction of on-treatment microbiota-modulating strategies. Fourth, the preservation of microbial diversity in the MBR-01 group was associated with a benefit in several domains related to QoL, suggesting a broader health benefit beyond diarrhea mitigation.

Overall, this pilot study demonstrates that MBR-01, a mixed probiotic/prebiotic intervention, may effectively reduce abemaciclib-induced diarrhea and preserve gut microbial diversity in patients with high-risk HR+/HER2– early breast cancer. A larger, randomized trial is warranted to confirm these results, to elucidate microbiota-mediated mechanisms, and to define the role of microbiota modulation in supportive oncology care. Future studies should also incorporate comprehensive longitudinal microbiota profiling, functional analyses, and biomarker discovery to clarify mechanistic pathways to potentially extend microbiota-based interventions beyond abemaciclib and optimize cancer therapy outcomes.

## Acknowledgements

We are grateful to all the patients and their families who participated in this study. Dr. F. Schettini is supported by a Juan Rodés clinician scientist contract from the Instituto de Salud Carlos III (JR24/00024).

## Data availability

The datasets generated and/or analyzed during the current study are available from the corresponding author upon reasonable request.

## Authors’ contributions

Dr. Generali conceived the study. Dr. Generali, Membrino, Gattazzo, Strina, Milani, De Bianco and Cervoni recruited and treated study participants. Dr. Schettini, Venturini, Castagnetti and Fontana performed the study analyses. All authors interpreted study results. Dr. Membrino, Generali, and Schettini wrote the first manuscript draft. All authors revised and approved the final manuscript version.

